# Longitudinal serologic and viral testing post–SARS-CoV-2 infection and post-receipt of mRNA COVID-19 vaccine in a nursing home cohort—Georgia, October 2020‒April 2021

**DOI:** 10.1101/2021.12.28.21268458

**Authors:** Farrell A. Tobolowsky, Michelle A. Waltenburg, Erin D. Moritz, Melia Haile, Juliana C. DaSilva, Amy J. Schuh, Natalie J. Thornburg, Adrianna Westbrook, Susannah L. McKay, Stephen P. LaVoie, Jennifer M. Folster, Jennifer L. Harcourt, Azaibi Tamin, Megan M. Stumpf, Lisa Mills, Brandi Freeman, Sandra Lester, Elizabeth Beshearse, Kristin D. Lecy, Laura G. Brown, Geroncio Fajardo, Jeanne Negley, L. Clifford McDonald, Preeta K. Kutty, Allison C. Brown, for the CDC Infection Prevention and Control Team

**Author notes:** **Corresponding Author:** Farrell A. Tobolowsky, DO, MS, Centers for Disease Control and Prevention, 1600 Clifton Road, Atlanta, GA, USA 30329. Both authors contributed equally to this work (Farrell A. Tobolowsky, Michelle A. Waltenburg). The findings and conclusions in this report are those of the authors and do not necessarily represent the official position of the Centers for Disease Control and Prevention. Use of trade names and commercial sources is for identification only and does not imply endorsement by the U.S. Department of Health and Human Services.

## Abstract

**Importance:** There are limited data describing SARS-CoV-2–specific immune responses and their durability following infection and vaccination in nursing home residents.

**Objective:** To evaluate the quantitative titers and durability of binding antibodies detected after SARS-CoV-2 infection and subsequent COVID-19 vaccination.

**Design:** A prospective longitudinal evaluation included nine visits over 150 days; visits included questionnaire administration, blood collection for serology, and paired anterior nasal specimen collection for testing by BinaxNOW™ COVID-19 Ag Card (BinaxNOW), reverse transcription polymerase chain reaction (RT-PCR), and viral culture.

**Setting:** A nursing home during and after a SARS-CoV-2 outbreak.

**Participants:** 11 consenting SARS-CoV-2–positive nursing home residents.

**Main Outcomes and Measures:** SARS-CoV-2 testing (BinaxNOW™, RT-PCR, viral culture); quantitative titers of binding SARS-CoV-2 antibodies post-infection and post-vaccination (beginning after the first dose of the primary series).

**Results:** Of 10 participants with post-infection serology results, 9 (90%) had detectable Pan-Ig, IgG, and IgA antibodies and 8 (80%) had detectable IgM antibodies. At first antibody detection post-infection, two-thirds (6/9, 67%) of participants were RT-PCR–positive but none were culture positive. Ten participants received vaccination; all had detectable Pan-Ig, IgG, and IgA antibodies through their final observation ≤90 days post-first dose. Post-vaccination geometric means of IgG titers were 10–200-fold higher than post-infection.

**Conclusions and Relevance:** Nursing home residents in this cohort mounted robust immune responses to SARS-CoV-2 post-infection and post-vaccination. The augmented antibody responses post-vaccination are potential indicators of enhanced protection that vaccination may confer on previously infected nursing home residents.

## Introduction

COVID-19 outbreaks in nursing homes have caused significant morbidity and mortality, including 740,034 cases and 141,084 deaths in residents as of December 5, 2021 (1). Although nursing home residents are typically older with multiple comorbidities compared with the general population, preliminary data show they frequently mount an immune response within 15 to 30 days of SARS-CoV-2 diagnosis (2, 3). However, the kinetics of SARS-CoV-2–specific binding antibody titers and isotypes that persist beyond the early acute phase of infection are unknown.

Vaccination is a key strategy to prevent COVID-19 morbidity and mortality in nursing home residents (4). Residents were prioritized for early COVID-19 vaccination because nursing homes experienced rapid transmission and severe disease outcomes. mRNA vaccines were the first available, and early post-emergency use authorization data demonstrated that the administration of mRNA vaccines led to a decrease in new SARS-CoV-2 infections in nursing home residents (5-7). However, in some instances, especially in the context of emerging SARS-CoV-2 variants, vaccinated residents have tested positive for SARS-CoV-2 and developed COVID-19 during routine screening and outbreaks (8, 9). Despite these infections in vaccinated persons, studies indicate that overall, mRNA vaccines have decreased COVID-19 morbidity and mortality in nursing home residents (10, 11).

Characterizing SARS-CoV-2–specific immune responses elicited through natural infection or vaccination, and their durability in nursing home residents, is critical to understanding potential immune protection against infection. While preliminary data show that nursing home residents mount initial immune responses to infection and vaccination, the duration and strength of this immune response remains unknown. Additionally, information on how antibody development relates to viral RNA detection and virus shedding in nursing home residents is limited. In a longitudinal evaluation of SARS-CoV-2–positive nursing home residents, we describe (a) how the clinical course of COVID-19 illness, severity of illness, and underlying medical conditions related to antibody development, (b) how repeated viral testing results correlated with antibody development post-diagnosis, and (c) the comparative titers and durability of immunoglobulin isotypes (i.e., Pan Ig, IgG, IgA, IgM) mounted in response to SARS-CoV-2 infection and receipt of the Pfizer-BioNTech COVID-19 Vaccine.

## Methods

A longitudinal evaluation to characterize SARS-CoV-2–specific immune responses was initiated in a 149-bed nursing home that began experiencing an outbreak of SARS-CoV-2 infections in October 2020. Residents were eligible to participate if they were medically stable, had decision-making capacity, and provided informed consent after testing SARS-CoV-2–positive using BinaxNOW™ COVID-19 Ag Cards (BinaxNOW) (Abbott; Scarborough, ME) in any of three rounds of Centers for Disease Control and Prevention (CDC)-led facility-wide point prevalence surveys (PPS) or by previous testing results generated by the facility (12, 13). The first facility-generated positive viral test occurred in early October 2020. CDC PPS occurred during late October to early November 2020 and the longitudinal evaluation included visits from October 25, 2020 through April 1, 2021.

Upon enrollment, the first four visits were conducted every other day. During these visits, CDC administered a standardized questionnaire to each participant, blood specimens were collected for serology, and paired bilateral anterior nasal specimens were collected for testing by BinaxNOW and real-time reverse transcription polymerase chain reaction (RT-PCR) (14). Participants were interviewed to obtain demographics and information about their COVID-19 exposures, signs, symptoms, and hospitalization. After the first four visits, follow-up visits occurred monthly for a total of nine visits (Figure 1). During these monthly follow-up visits, one nasal specimen was collected for RT-PCR, blood specimens were collected for serology, and participants were interviewed about COVID-19 signs, symptoms, and hospitalizations since the previous visit.

**Figure 1.**
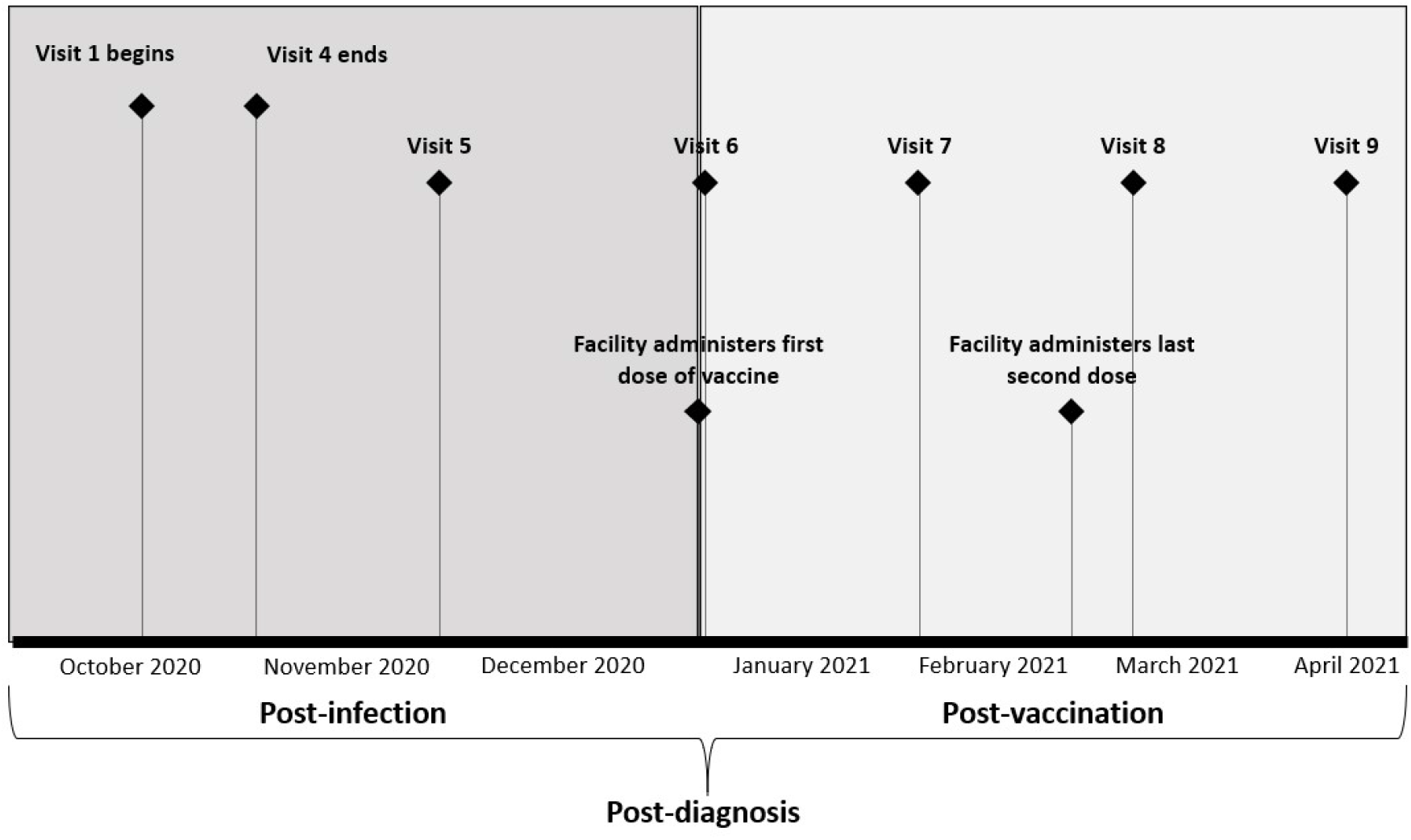
Timeline of participant visits and evaluation activities conducted, by time period—Georgia. October 2020‒April 2021 Upon enrollment after a SARS-CoV-2–positive diagnosis by BinaxNOW™ COVID-19 Ag Cards and/or real-time reverse transcription polymerase chain reaction, the first four visits were conducted every other day. Visits 5–9 occurred monthly. The post-diagnosis period was defined as the time after each participant’s first positive SARS-CoV-2 test result. The post-infection period was defined as the time from SARS-CoV-2 diagnosis to receipt of first Pfizer-BioNTech COVID-19 Vaccine dose. The post-vaccination period was defined as the time after receipt of first vaccine dose to the end of the evaluation period. Nine participants received their two vaccine doses 21 days apart; one participant received their two vaccine doses 28 days apart.

The facility began administering the primary series of mRNA COVID-19 vaccines during the evaluation period (4); willing participants were vaccinated between January–February 2021. Timing of vaccination was determined by the facility and timing of receipt of the first vaccine dose varied by participant. CDC visits 6–9 occurred after facility vaccine clinics were initiated. In this evaluation, participants were considered fully vaccinated two weeks after receiving the second dose of the mRNA COVID-19 vaccine.

Electronic medical chart abstraction was performed to obtain supplemental information on participants’ past medical history, medications, laboratory test results, clinical outcomes, and vaccine administration. Information from questionnaires and chart abstraction were entered into a Research Electronic Data Capture (REDCap) database (15, 16). If visits occurred while a participant was hospitalized, specimens and clinical data were not collected from that participant; however, that participant could choose to continue in the evaluation upon return to the nursing home.

Reported COVID-19 symptom status was characterized as asymptomatic or symptomatic, and participants’ illnesses were further characterized as mild, moderate, or severe. A participant was asymptomatic if he or she did not report any COVID-19 signs or symptoms in the 14 days before testing positive or during the evaluation period. Mild illness included participants with asymptomatic COVID-19 infections and those who reported COVID-19 signs or symptoms without dyspnea or evidence of lower respiratory tract involvement. Moderate illness included participants with signs of lower respiratory disease by clinical assessment or imaging, and severe disease included participants with tachypnea (>30 breaths per minute), oxygen saturation <90% (or decrease from baseline over 3% for at least two consecutive values), hospitalization, or death (17). An immunocompromised condition included the following: recent or active malignancy, bone marrow transplant, solid organ transplant, primary or secondary immune deficiency, or the use of oral or intravenous steroids or any immunosuppressant drugs.

Details of specimen collection, BinaxNOW, RT-PCR, and viral culture methods have been described previously (12). Viral culture was conducted on RT-PCR–positive specimens with a cycle threshold (Ct) value <35. Serum specimens were tested for anti–SARS-CoV-2 antibodies using an enzyme-linked immunosorbent assay targeting the extracellular domain of the SARS-CoV-2 spike protein (18). This assay uses anti–pan-immunoglobulin (Ig) secondary antibodies that detect any SARS-CoV-2 immunoglobulin isotype, including IgG, IgM, and IgA. Seroconversion was defined as a signal to threshold >1 at a serum dilution of 1:100 for any isotype; seroreversion occurred when a previously seropositive participant became seronegative.

Day 0 for each participant was defined as the day of their first SARS-CoV-2–positive test (BinaxNOW or RT-PCR) by either the facility or CDC PPS. We defined the entire evaluation period as “post-diagnosis”, the time period after each participant’s SARS-CoV-2 diagnosis and before vaccination as “post-infection,” and the time period after a participant received their first vaccine dose of the primary series as “post-vaccination” (Figure 1).

Geometric means of serum antibody titers were calculated among all participants with specimens collected during specific post-infection and post-vaccination time periods: 0–14, 15–30, 31– 60, and 61–90 days. Ratios for mean titers post-vaccination to mean titers post-infection were calculated for each isotype; only antibody titers >100 were included in these ratios. Median serum antibody titers and ratios of median titers post-vaccination to post-infection were also calculated. Data were analyzed by Microsoft Excel (Microsoft Office 365 Version; Redmond, WA) and SAS, version 9.4 (SAS Institute; Cary, NC).

This evaluation was reviewed by the National Center for Emerging and Zoonotic Infectious Diseases Human Studies Coordinator at the CDC and was determined to be non-research public health surveillance as defined in 45 CF 46.102 (19-23). It was determined that ethical review by the CDC Human Research Protection Office and IRB review was not required. This work did not receive any non-CDC funding support.

## Results

### Demographic and clinical characteristics

Among 127 residents who participated in at least one CDC PPS, 47 (37%) antigen-positive residents were identified; 31 (66%) antigen-positive residents were eligible for enrollment in this longitudinal evaluation, of which 12 (39%) agreed to participate. Eight (67%) participants tested positive by facility testing conducted prior to CDC PPS. One participant died before the second follow-up visit and was excluded from analysis. The median age of the remaining 11 participants was 74 years (range: 37– 90 years) and seven (64%) were male (Table 1). Seven (64%) participants were White and four (36%) were Black; no participants reported Hispanic ethnicity. The most common underlying conditions were cardiovascular disease (11, 100%) and diabetes (6, 55%); 10 (91%) participants had ≥3 underlying conditions (Table 1). One participant (A) was immunocompromised with B-cell lymphoma. Four (36%) participants (C, D, G, H) were classified as having severe COVID-19 illness, all of whom required hospitalization; one (D) died during follow-up after visit 8. Two (18%) participants (E, J) were asymptomatic. Ten of 11 (91%) participants received two doses of the Pfizer-BioNTech COVID-19 Vaccine during the evaluation period; the other participant (C) refused the vaccine.

**Table 1.**
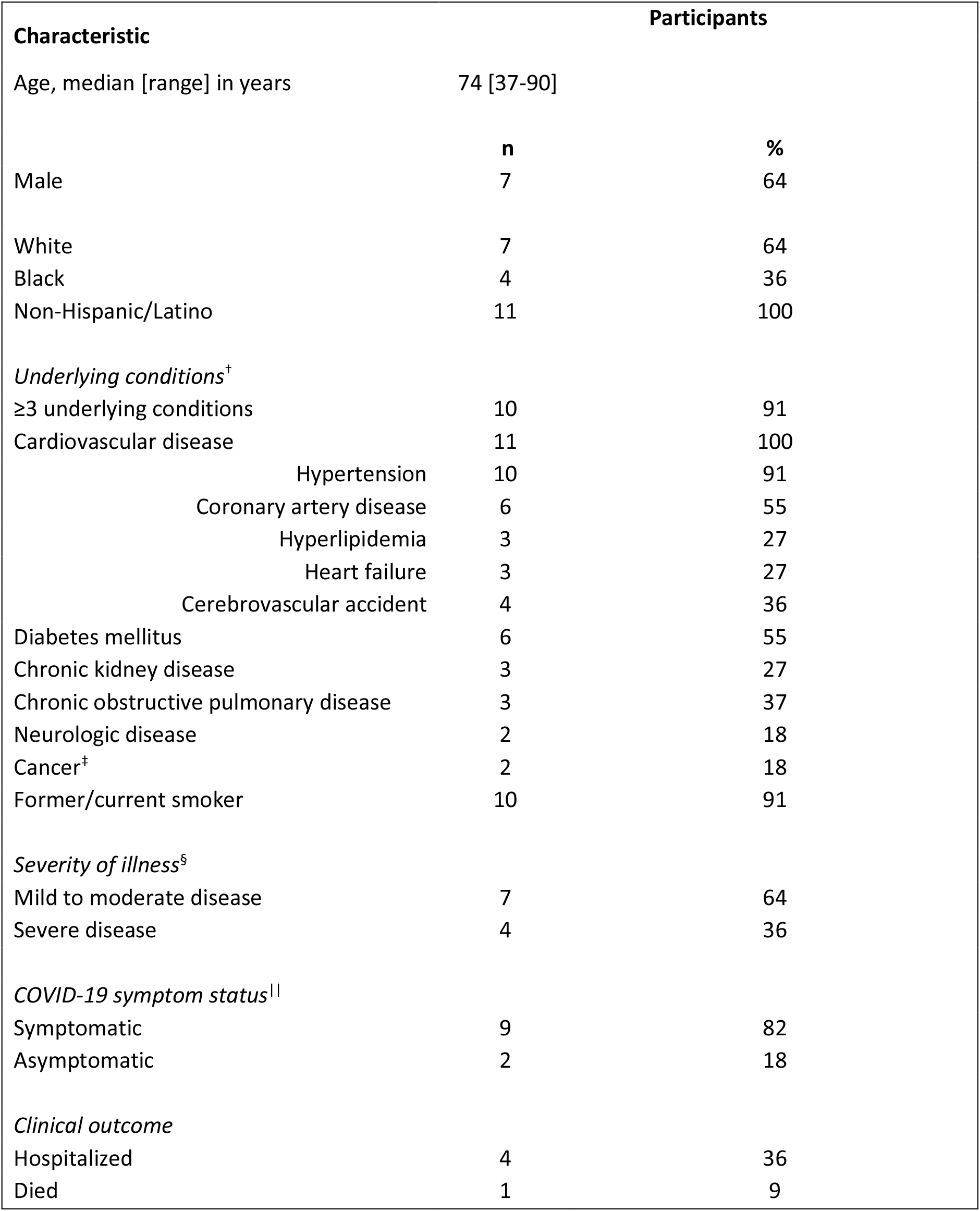
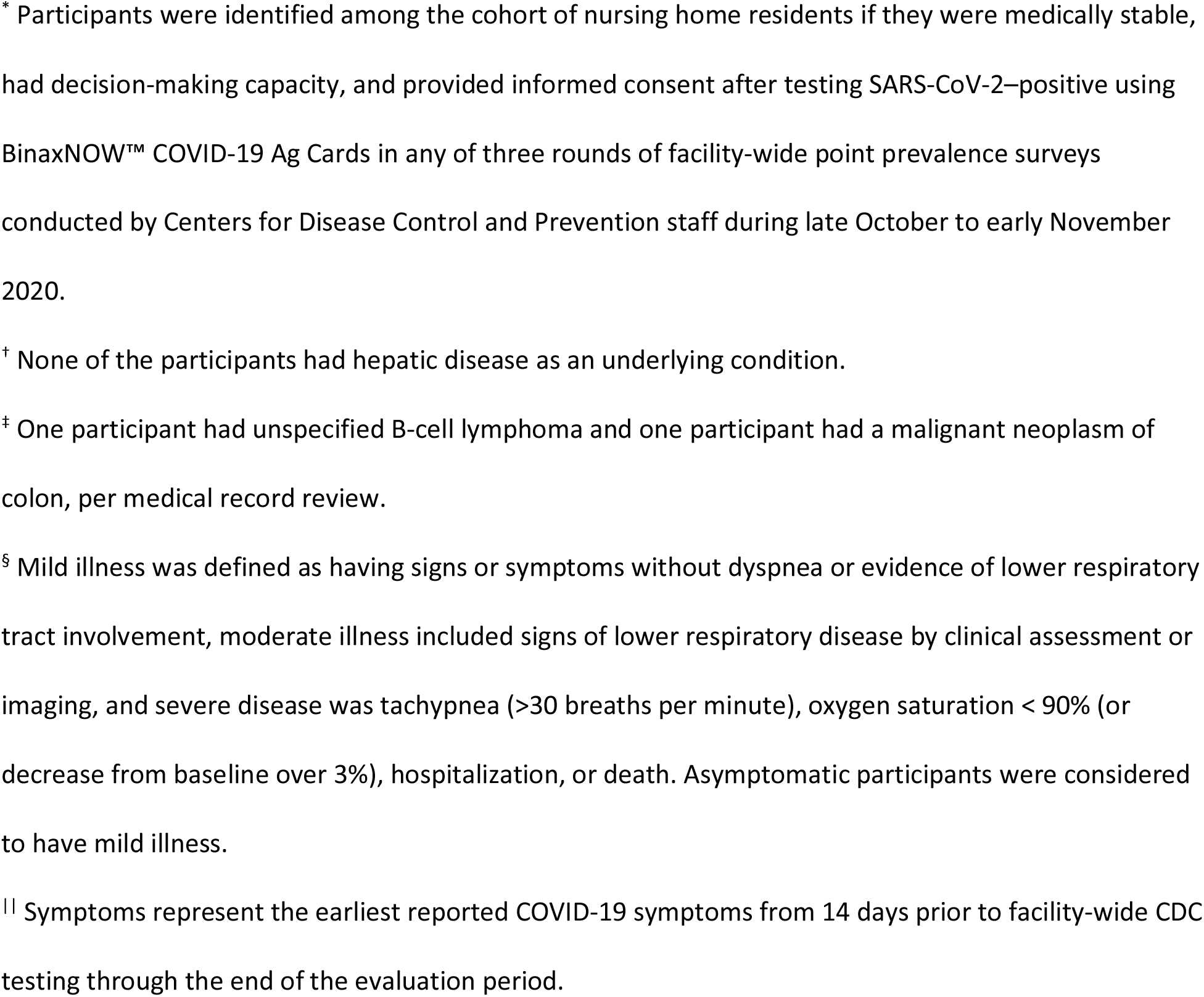
Demographic and clinical characteristics of participants^*^ (N=11) in a nursing home cohort— Georgia, October 2020‒April 2021.

| Characteristic |  | Participants |
| --- | --- | --- |
| Age, median [range] in years | 74 [37-90] |  |
|  | **n** | **%** |
| Male | 7 | 64 |
| White | 7 | 64 |
| Black | 4 | 36 |
| Non-Hispanic/Latino | 11 | 100 |
| *Underlying conditions<sup>†</sup>* |  |  |
| ≥3 underlying conditions | 10 | 91 |
| Cardiovascular disease | 11 | 100 |
| Hypertension | 10 | 91 |
| Coronary artery disease | 6 | 55 |
| Hyperlipidemia | 3 | 27 |
| Heart failure | 3 | 27 |
| Cerebrovascular accident | 4 | 36 |
| Diabetes mellitus | 6 | 55 |
| Chronic kidney disease | 3 | 27 |
| Chronic obstructive pulmonary disease | 3 | 37 |
| Neurologic disease | 2 | 18 |
| Cancer<sup>‡</sup> | 2 | 18 |
| Former/current smoker | 10 | 91 |
| *Severity of illness<sup>§</sup>* |  |  |
| Mild to moderate disease | 7 | 64 |
| Severe disease | 4 | 36 |
| *COVID-19 symptom status* |  |  |
| Symptomatic | 9 | 82 |
| Asymptomatic | 2 | 18 |
| *Clinical outcome* |  |  |
| Hospitalized | 4 | 36 |
| Died | 1 | 9 |

\* Participants were identified among the cohort of nursing home residents if they were medically stable, had decision-making capacity, and provided informed consent after testing SARS-CoV-2–positive using BinaxNOW™ COVID-19 Ag Cards in any of three rounds of facility-wide point prevalence surveys conducted by Centers for Disease Control and Prevention staff during late October to early November 2020.
† None of the participants had hepatic disease as an underlying condition.
‡ One participant had unspecified B-cell lymphoma and one participant had a malignant neoplasm of colon, per medical record review.
§ Mild illness was defined as having signs or symptoms without dyspnea or evidence of lower respiratory tract involvement, moderate illness included signs of lower respiratory disease by clinical assessment or imaging, and severe disease was tachypnea (>30 breaths per minute), oxygen saturation < 90% (or decrease from baseline over 3%), hospitalization, or death. Asymptomatic participants were considered to have mild illness.
|| Symptoms represent the earliest reported COVID-19 symptoms from 14 days prior to facility-wide CDC testing through the end of the evaluation period.

### Laboratory results

A total of 166 anterior nasal specimens (53 paired for RT-PCR and BinaxNOW; 60 unpaired for RT-PCR only) and 61 blood specimens were analyzed. Participants had a median of 6 (interquartile range [IQR]: 5–7) blood specimens collected across a median of 141 days (IQR: 141–163). One participant (G) did not provide blood specimens during the post-infection period but did during the post-vaccination period. All blood specimens from participant C were considered post-infection due to vaccination refusal.

### Antibody profile post-infection and prior to vaccination

Of 10 participants with post-infection serology results, 9 (90%) had detectable Pan-Ig, IgG, and IgA antibodies, 8 (80%) had detectable IgM antibodies, and one participant (I) did not mount any binding antibodies post-infection. Among the seven participants with serology conducted within 30 days post-infection, the median time to detection was 13 days for Pan-Ig and IgG (IQR: 11–17); 16 days for IgA (IQR: 12–17). Six (86%) of these participants with IgM antibodies had a median time to detection of 15 days (IQR: 12–17).

At the time of first antibody detection post-infection, two-thirds (6/9, 67%) of participants were still RT-PCR–positive but none were viral culture positive (Appendix Figure 1). The median Ct value at the time of first antibody detection for these participants was 29 (IQR: 25–34). Participant G did not have blood specimens collected post-infection and participant I did not have detectable antibodies post-infection, but both were RT-PCR–negative and had prior negative viral culture results at the time of first antibody detection. Antibody detection was either concurrent with or subsequent to negative viral culture results but did not correlate with negative RT-PCR or BinaxNOW results (Appendix Figure 1).

Pan-Ig and IgG rose and remained elevated post-infection (Figure 2). IgM and IgA titers started to decline during the 31–60 days post-infection; seroreversion in these isotypes occurred in some participants (J, K, L) by 50 days post-infection and before vaccination (Appendix Figure 2). The participant (C) who declined vaccination had detectable Pan-Ig, IgG, and IgA antibodies for at least 150 days post-diagnosis (Appendix Figure 3). Participant H had lower post-infection antibody titers overall and had lymphopenia as determined in a blood specimen collected within 30 days prior to enrollment.

**Figure 2.**
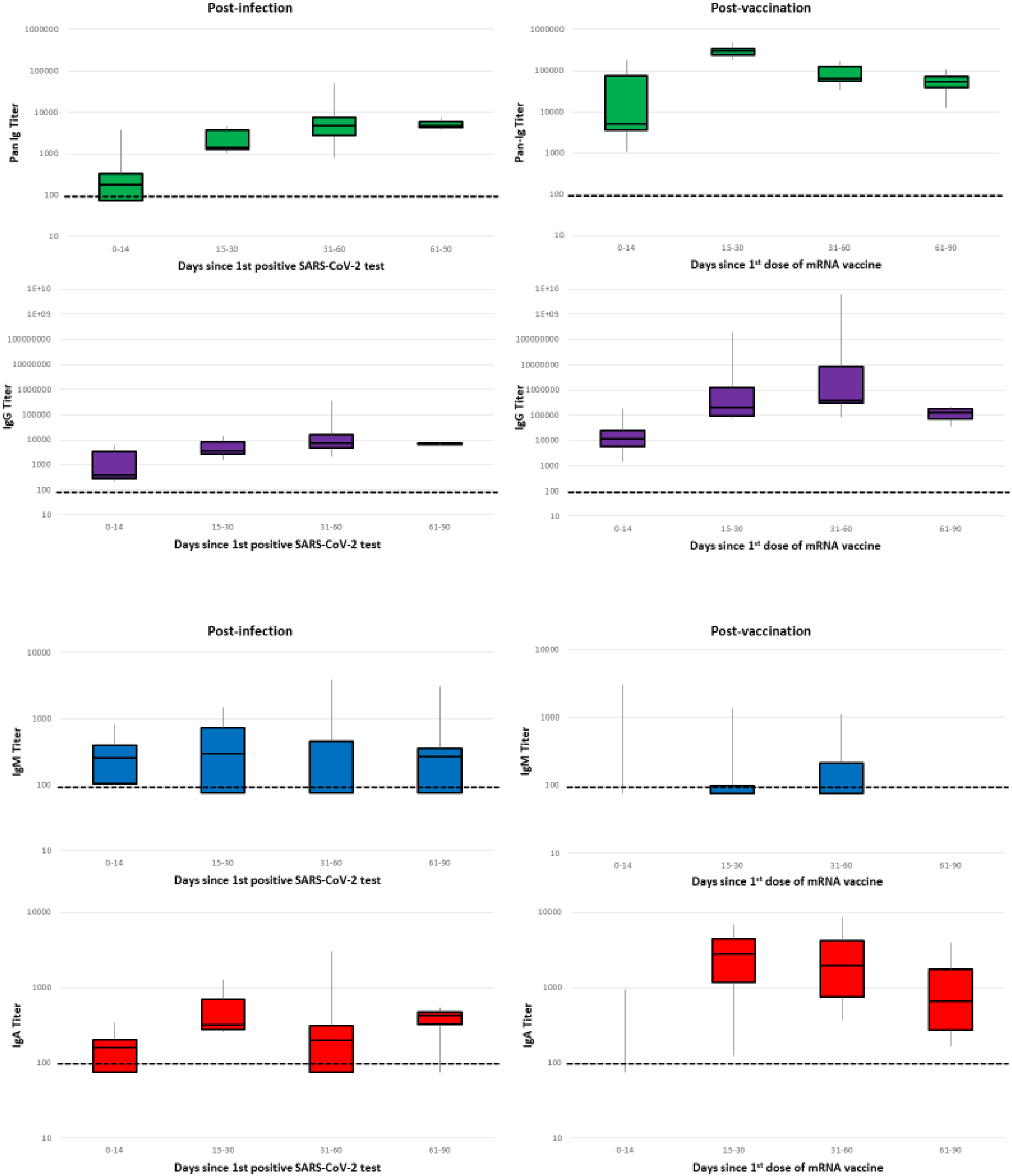
Distribution of anti–SARS-CoV-2 serum antibody titers by isotype and time period among participants (N=11) in a nursing home cohort—Georgia, October 2020‒April 2021. The post-infection period was defined as the time from SARS-CoV-2 diagnosis (by BinaxNOW™ COVID-19 Ag Cards and/or real-time reverse transcription polymerase chain reaction) to receipt of first Pfizer-BioNTech COVID-19 Vaccine dose. The post-vaccination period was defined as the time after receipt of first vaccine dose to the end of the evaluation period. Participant G was not included in the post-infection analyses because they did not provide blood specimens during this time period. Participant C was not included in the post-vaccination analyses due to vaccination refusal. Participants with serum antibody titers below the seroconversion threshold (defined as a signal threshold >1 at the 1:100 dilution for any isotype) were assigned a value of 25 for graphical representation only. In each box and whisker plot, the horizontal line represents the median, and the top and bottom of the box the interquartile range, and the whiskers the minimum and maximum values. The y-axis is plotted in logarithmic scale. Only three observations were above the limit of detection during 0–14 days post-vaccination for IgM and IgA; IgM titers during this time period ranged from 313–3013 and IgA titers ranged from 177–890.

### Antibody profile post-vaccination

Among the 10 participants vaccinated during the evaluation period, timing of the first vaccine dose varied by participant and occurred between 71–105 days post-diagnosis. All 10 (100%) had detectable Pan-Ig, IgG, and IgA antibodies and six (60%) had detectable IgM antibodies post-vaccination (Appendix Figure 2). Pan-Ig, IgG, and IgA titers began to decrease during 61–90 days post-vaccination (141 days post-diagnosis) (Figure 2).

Post-infection geometric means were highest during 15–30 days for IgA, 31–60 days for IgM and IgG, and 61–90 days for Pan-Ig (Table 2). Post-vaccination geometric means were highest during 0–14 days for IgM, 15–30 days for Pan-Ig, and 61–90 days for IgG and IgA (Table 2). Participants demonstrated higher Pan-Ig, IgG, and IgA titers post-second vaccine dose than pre-first dose. The largest geometric mean titer ratios were measured during specific post-vaccination and post-infection time periods: 15–30 days for Pan-Ig (142.3) and 31–60 days for IgG (200.5) and IgA (4.4). Geometric mean IgM titer ratios were not calculated during 61–90 days because all post-vaccination titers were below the limit of detection. Median serum antibody titers and ratios during post-infection and post-vaccination time periods are displayed in Appendix Table 1.

**Table 2.**
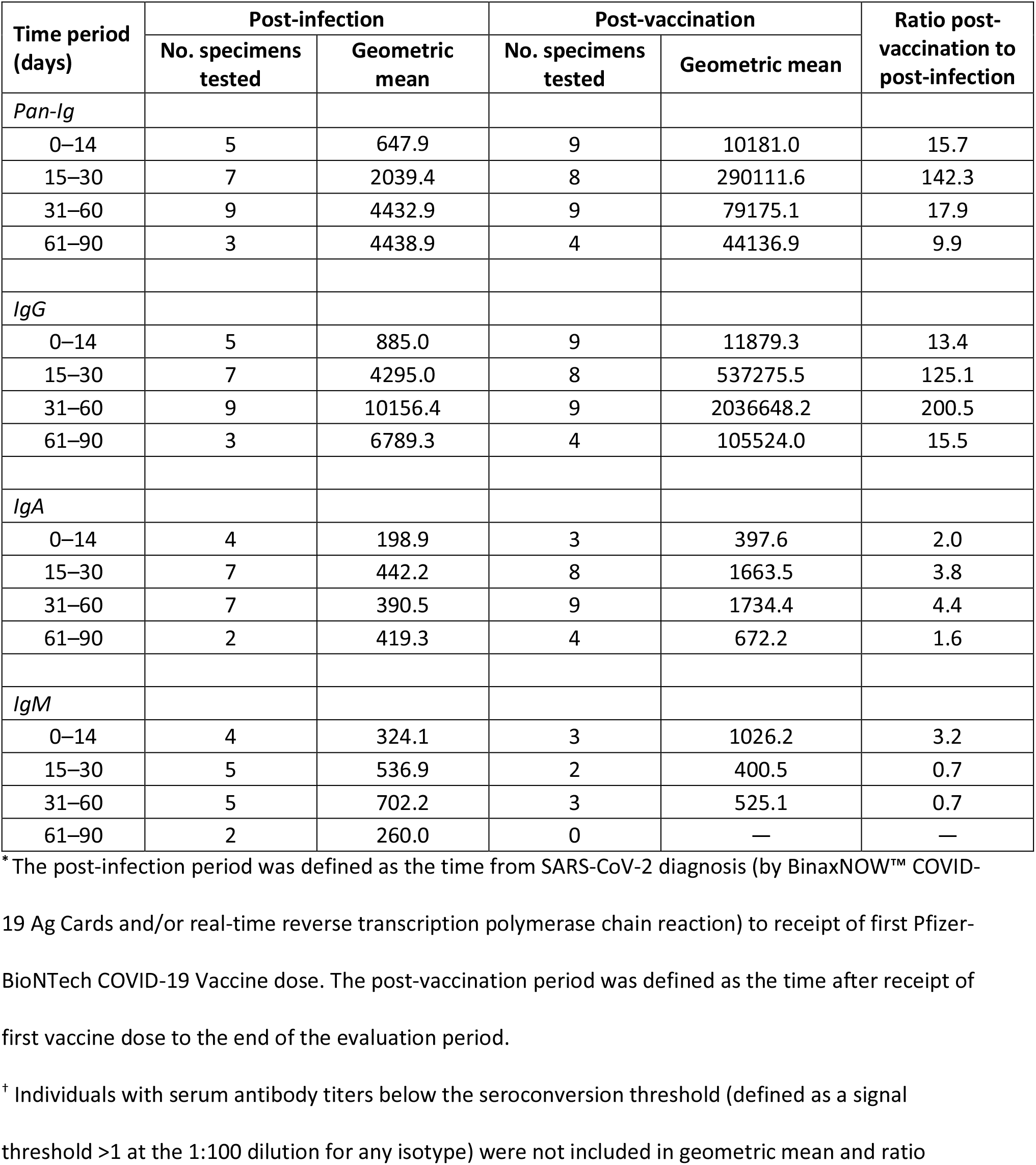
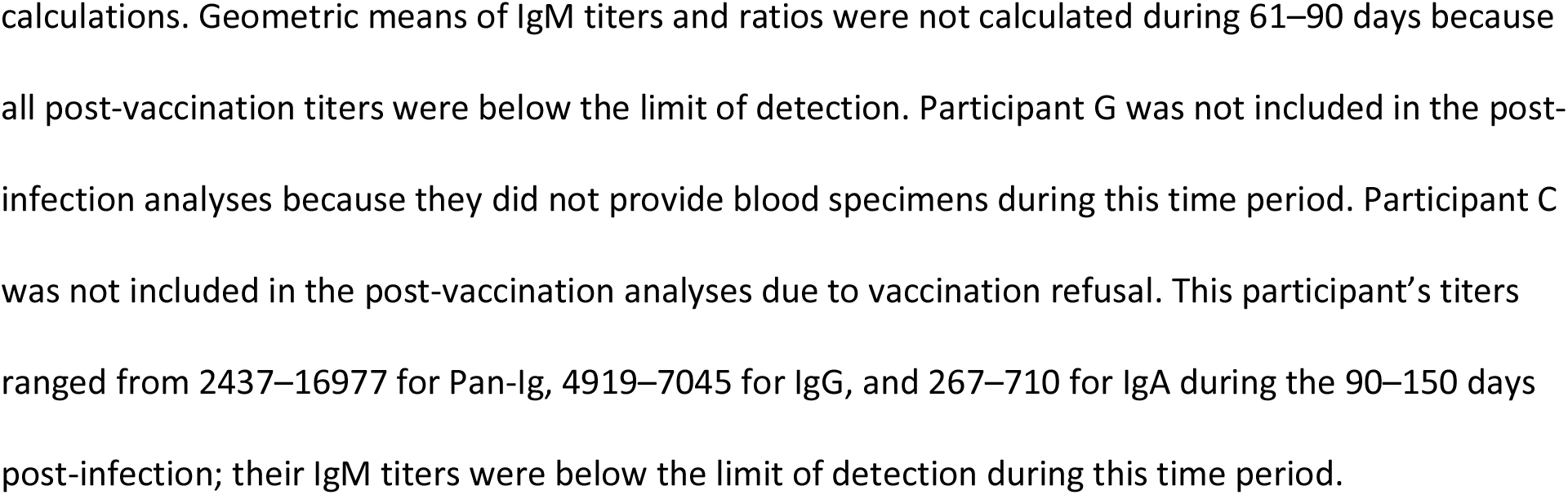
Geometric means of anti–SARS-CoV-2 serum antibody titers and ratios post-vaccination and post-infection by isotype among participants (N=11) in a nursing home cohort—Georgia, October 2020‒April 2021*,†.

| Time period (days) | Post-infection |  | Post-vaccination |  | Ratio post-vaccination to post-infection |
| --- | --- | --- | --- | --- | --- |
|  | No. specimens tested | Geometric mean | No. specimens tested | Geometric mean |  |
| *Pan-Ig* |  |  |  |  |  |
| 0–14 | 5 | 647.9 | 9 | 10181.0 | 15.7 |
| 15–30 | 7 | 2039.4 | 8 | 290111.6 | 142.3 |
| 31–60 | 9 | 4432.9 | 9 | 79175.1 | 17.9 |
| 61–90 | 3 | 4438.9 | 4 | 44136.9 | 9.9 |
| *IgG* |  |  |  |  |  |
| 0–14 | 5 | 885.0 | 9 | 11879.3 | 13.4 |
| 15–30 | 7 | 4295.0 | 8 | 537275.5 | 125.1 |
| 31–60 | 9 | 10156.4 | 9 | 2036648.2 | 200.5 |
| 61–90 | 3 | 6789.3 | 4 | 105524.0 | 15.5 |
| *IgA* |  |  |  |  |  |
| 0–14 | 4 | 198.9 | 3 | 397.6 | 2.0 |
| 15–30 | 7 | 442.2 | 8 | 1663.5 | 3.8 |
| 31–60 | 7 | 390.5 | 9 | 1734.4 | 4.4 |
| 61–90 | 2 | 419.3 | 4 | 672.2 | 1.6 |
| *IgM* |  |  |  |  |  |
| 0–14 | 4 | 324.1 | 3 | 1026.2 | 3.2 |
| 15–30 | 5 | 536.9 | 2 | 400.5 | 0.7 |
| 31–60 | 5 | 702.2 | 3 | 525.1 | 0.7 |
| 61–90 | 2 | 260.0 | 0 | — | — |

<sup>\*</sup> The post-infection period was defined as the time from SARS-CoV-2 diagnosis (by BinaxNOW™ COVID-19 Ag Cards and/or real-time reverse transcription polymerase chain reaction) to receipt of first Pfizer-BioNTech COVID-19 Vaccine dose. The post-vaccination period was defined as the time after receipt of first vaccine dose to the end of the evaluation period.
<sup>†</sup> Individuals with serum antibody titers below the seroconversion threshold (defined as a signal threshold >1 at the 1:100 dilution for any isotype) were not included in geometric mean and ratio
calculations. Geometric means of IgM titers and ratios were not calculated during 61–90 days because all post-vaccination titers were below the limit of detection. Participant G was not included in the post- infection analyses because they did not provide blood specimens during this time period. Participant C was not included in the post-vaccination analyses due to vaccination refusal. This participant's titers ranged from 2437–16977 for Pan-Ig, 4919–7045 for IgG, and 267–710 for IgA during the 90–150 days post-infection; their IgM titers were below the limit of detection during this time period.

Participant J had a substantial increase in IgG titers post-vaccination; this participant received their first vaccine dose on day 105 post-diagnosis, much later than most participants who received their first vaccine dose earlier (Appendix Figure 3). Participant A who was immunocompromised with B-cell lymphoma and classified as having mild to moderate COVID-19 disease had similar titers as other participants. The unvaccinated participant (C) had lower Pan-Ig and IgG titers than vaccinated participants after 90 days post-diagnosis (Appendix Figure 2).

## Discussion

Although there have been concerns about immunosenescence causing an inadequate immune response to SARS-CoV-2 infection and vaccination in the nursing home population, anti–SARS-CoV-2 antibodies were consistently detected throughout the evaluation period in this nursing home cohort. Furthermore, antibody responses in previously infected participants were augmented after vaccination and were still present throughout the evaluation period.

Some participants were antigen and RT-PCR positive at the time of first antibody detection; however, no culturable virus was identified in respiratory specimens following detection of antibodies. In general population studies, infected individuals may initially have positive nucleic acid amplification tests (including RT-PCR) when anti–SARS-CoV-2 antibodies are first detected but are markedly less likely to have positive results following antibody development (24). Specifically, detection of binding IgG antibodies is associated with the absence of replication-competent virus (25). As a positive viral culture is a likely indicator of the presence of replication-competent virus, our findings further support seroconversion as a potential marker of non-infectivity post-infection and pre-vaccination (26).

Most participants developed all anti–SARS-CoV-2 antibody isotypes that were tested post-infection, highlighting the diversity of antibody development in this nursing home cohort. The participant who did not develop antibodies post-infection was only tested on Days 3 and 8. Pan-Ig and IgG titers rose and remained elevated post-infection while IgM and IgA titers began to dissipate around 30 days post-infection. The duration of IgA positivity was longer than IgM. Similar kinetics have been described in previous studies (27, 28). Prior reports have also suggested that higher titers are more commonly associated with severe disease (29-31). In this cohort, quantitative antibody development did not differ between those with mild to moderate and severe illness; however, our small sample size may have limited our ability to detect a meaningful difference.

Prior studies reported that post-infection antibodies can be detected for at least six months after diagnosis in the general population (32) and in nursing home cohorts (33). In this assessment, one unvaccinated participant (C) had Pan-Ig, IgG, and IgA antibodies detected at 150 days (five months) post-infection when our evaluation period ended, which may indicate antibody durability even in the absence of vaccination. While our findings add to the growing body of evidence that nursing home residents can maintain detectable binding antibodies for at least five months after diagnosis of SARS-CoV-2 infection, we were unable to determine the maximum duration of post-infection antibodies because most participants in our cohort were vaccinated 71–105 days post-diagnosis.

This analysis highlights the augmented impact of COVID-19 vaccination on post-infection antibody titers, and likely enhanced immune protection, in previously infected nursing home residents (34, 35). Evaluating titers of specific binding isotypes, participants had more robust Pan-Ig, IgG, and IgA titers post-vaccination. Post-vaccination geometric mean titers of IgG were 10–200-fold higher than post-infection, which is consistent with reported post-vaccination titer increases in healthcare workers who had previous SARS-CoV-2 infection (36, 37). While more information is needed to understand the correlation of post-mRNA vaccination titers with immune protection, a study by Feng et al. among recipients of the ChAdOx1 nCoV-19 vaccine demonstrated a correlation between binding IgG antibody titers and vaccine efficacy (38). Moreover, Feng et al. were able to derive threshold antibody levels that correlated with differing levels of vaccine efficacy against symptomatic SARS-CoV-2 infection (38). Applying these criteria to our nursing home cohort, all vaccinated participants surpassed the IgG titer threshold that correlated with 80% vaccine efficacy two weeks after receiving their second vaccine dose.

Previous studies have shown increases in both IgG and IgA post-vaccination, with IgA detected in both serum and mucosal sources (39-41). Robust IgG and IgA responses likely induce systemic and mucosal protection (42). IgA has been found to dominate the early neutralizing antibody response to SARS-CoV-2; a similar IgA response after mRNA vaccination has been hypothesized (42). In this evaluation, post-vaccination geometric mean titers of IgA were 2–4-fold higher than post-infection titers, suggesting that IgA could be used as an indicator of mRNA vaccine response.

Our evaluation had several limitations. We had a small sample size, preventing statistical comparisons and potentially limiting the generalizability of our findings. Participants enrolled in our evaluation only received the Pfizer-BioNTech COVID-19 Vaccine; therefore, we were unable to describe the impact of vaccination with other products. Some participants were unable to provide blood specimens when requested and most visits occurred only monthly, so we likely missed the exact timing of seroconversion, peak antibody titers, and seroreversion. Not detecting antibodies in one participant during this first 30 days post-infection was possibly an artifact of timing our collections relative to their infection. Descriptions of antibody duration were limited by the length of the evaluation period. Seven participants had visits that occurred one day after their first vaccine; though these visits were categorized as post-vaccination, we do not know if titers observed from this visit were a continuation of responses primed by their previous infection or a result of the vaccine already augmenting that response. Therefore, we may have underestimated the scale of post-vaccination augmentation because those titer calculations were influenced by values from measurements that were likely more reflective of the post-infection time period. Limited electronic medical record documentation of vital signs, including oxygen saturation, and the difficulty of discerning COVID-19 symptomology in a nursing home cohort with multiple underlying comorbidities, chronic symptoms, and a modified clinical presentation of particular signs and symptoms made it challenging to classify severity of illness. Lastly, since this evaluation occurred before the rise of the SARS-CoV-2 B.1.617.2 (Delta) and B.1.1.529 (Omicron) variants in the United States, we do not know how participants’ post-infection and post-vaccination antibody responses could differ in the setting of these and other emerging SARS-CoV-2 variants.

## Conclusion

The duration and diversity of binding antibodies observed in this cohort of nursing home residents could have implications for durability of immunity to SARS-CoV-2. Our findings of augmented Pan-Ig, IgG, and IgA antibody responses post-vaccination in nursing home residents with recent infection underscore the importance of vaccination efforts, especially where there is vaccine hesitancy among nursing home staff (43). While it is unknown how long these anti–SARS-CoV-2 antibodies persist following vaccination, findings from this evaluation suggest that nursing home residents do mount a robust humoral immune response to SARS-CoV-2 post-infection and post-vaccination.

Members of the CDC Infection Prevention and Control Team who contributed to this work but did not author it: Amelia Bhatnagar, BS; Jonathan Bryant-Genevier, PhD; Dustin W. Currie, PhD, MPH; Davina Campbell, MS, MPH; Sarah E. Gilbert, MPH; Kelly M. Hatfield, MSPH; David A. Jackson, MD, MPH; John A. Jernigan, MD, MS; James L. Dawson, PhD; Matthew J. Hudson, MD, MPH; Kahaliah Joseph, MSc; Sujan C. Reddy, MD, MSc; and Malania M. Wilson, MS, MBA.

## Data Availability

Data produced in the present study are available upon reasonable request to the authors.

## Acknowledgements

We are grateful to all residents and staff at the nursing home for their participation and support of this evaluation during an especially challenging time. We also thank the Georgia Department of Public Health.

