## Supplemental Material for "Longitudinal serologic and viral testing post–SARS-CoV-2 infection and post-receipt of mRNA COVID-19 vaccine in a nursing home cohort—Georgia, October 2020‒April 2021"

**Appendix Table 1. Median anti–SARS-CoV-2 serum antibody titers and ratios post-vaccination and post-infection by isotype among participants (N=11) in a nursing home cohort—Georgia, October 2020–April 2021[*]**

| Time period (days) | Post-infection | | Post-vaccination | | Ratio post-vaccination to post-infection |
|---|---|---|---|---|---|
| | No. specimens tested | Median titer (IQR) [†] | No. specimens tested | Median titer (IQR)[‡§] | |
| *Pan-Ig* | | | | | |
| 0–14 | 5 | 286 (257–1803) | 9 | 5137 (3584–73070) | 18.0 |
| 15–30 | 7 | 1,441 (1289–3682) | 8 | 299,721 (240573–345136) | 208.0 |
| 31–60 | 9 | 4,769 (2832–7582) | 9 | 64,344 (55171–127753) | 13.5 |
| 61–90 | 3 | 4,763 (2832–6153) | 4 | 53,664 (38454–71967) | 11.3 |
| *IgG* | | | | | |
| 0–14 | 5 | 395 (298–3340) | 9 | 11,758 (5797–25900) | 29.8 |
| 15–30 | 7 | 3,563 (2662–8028) | 8 | 206,062 (95411–1258378) | 57.8 |
| 31–60 | 9 | 7,472 (4952–15971) | 9 | 380,248 (318681–8705015) | 50.9 |
| 61–90 | 3 | 6,973 (6495–7217) | 4 | 130,964 (73799–185779) | 18.8 |
| *IgA* | | | | | |
| 0–14 | 3 | 155 (134–223) | 3 | 399 (288–645) | 2.6 |
| 15–30 | 7 | 318 (277–697) | 8 | 2,777 (1178–4462) | 8.7 |
| 31–60 | 7 | 198 (154–1255) | 9 | 1,956 (754–4242) | 9.9 |
| 61–90 | 3 | 399 (379–445) | 4 | 656 (272–1742) | 1.6 |

Abbreviations: IQR = interquartile range

[*] The post-infection period was defined as the time from SARS-CoV-2 diagnosis (by BinaxNOW™ COVID-19 Ag Cards and/or real-time reverse transcription polymerase chain reaction) to receipt of first Pfizer-BioNTech COVID-19 Vaccine dose. The post-vaccination period was defined as the time after receipt of first vaccine dose to the end of the evaluation period. Individuals with serum antibody titers below the seroconversion threshold (defined as a signal threshold >1 at the 1:100 dilution for any isotype) were not included in median and ratio calculations.

[†] Median post-infection titers peaked during 15–30 days for IgM, 31–60 days for Pan-Ig and IgG, and 61–90 days for IgA.

[‡] Participant C was not included in the post-vaccination analyses due to vaccination refusal. This participant’s titers ranged from 2437–16977 for Pan-Ig, 4919–7045 for IgG, and 267–710 for IgA during the 90–150 days post-infection; their IgM titers were below the limit of detection during this time period.

[§] Median post-vaccination titers peaked during 15–30 days for Pan-Ig and IgA, and 31–60 days for IgG. Post-vaccination median IgM titers were not calculated because most post-vaccination titers were below the limit of detection.

**Appendix Figure 1. Detection of SARS-CoV-2 by BinaxNOW™ rapid antigen test, real-time reverse transcription polymerase chain reaction, and viral culture by Pan-Ig serum antibody titers and days post-diagnosis of participants (N=11) in a nursing home cohort—Georgia, October 2020–April 2021**

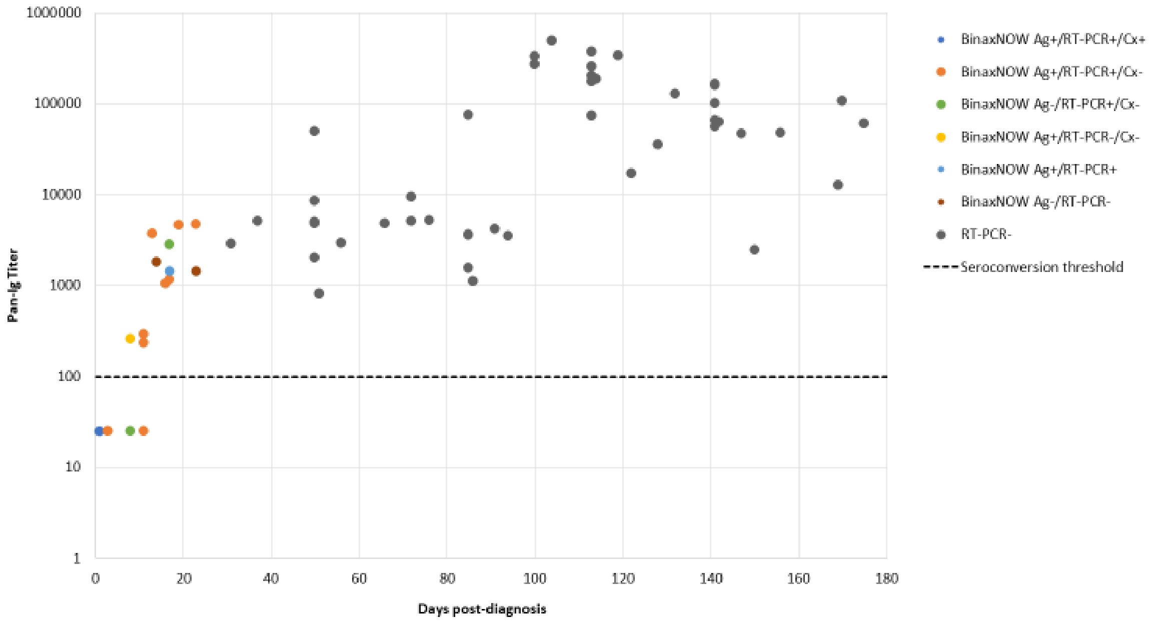

Abbreviations: BinaxNOW = BinaxNOW™ COVID-19 Ag Card; RT-PCR = real-time reverse transcription polymerase chain reaction; Cx = viral culture

Blood samples were obtained at several timepoints between 1–150 days post-diagnosis (time period after each participant's first positive SARS-CoV-2 test result). Due to challenges phlebotomizing some participants, serum titers were not generated for each participant for each visit. Participants with serum antibody titers below the seroconversion threshold (defined as a signal threshold >1 at the 1:100 dilution for any isotype) were assigned a value of 25 for graphical representation only. The y-axis is plotted in logarithmic scale.

**Appendix Figure 2. Anti–SARS-CoV-2 serum antibody titers by isotype and days post-diagnosis among participants (N=11) in a nursing home cohort—Georgia, October 2020–April 2021**

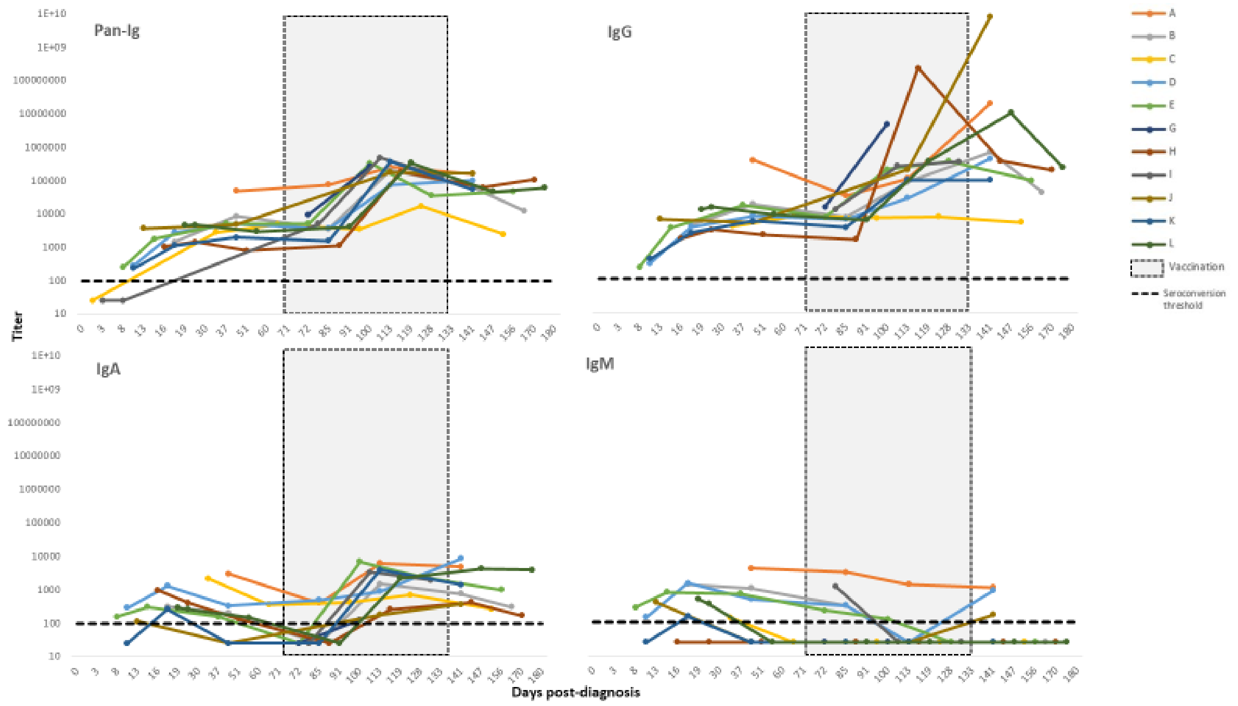

Colored lines represent individual participants. Blood samples were obtained at several timepoints between 1–150 days post-diagnosis (time period after each participant's first positive SARS-CoV-2 test result). Due to challenges phlebotomizing some patients, serum titers were not generated for each participant for each visit. Pfizer-BioNTech COVID-19 vaccines were administered at the facility between 71–105 days post-diagnosis; participant C declined vaccination. Participants with serum antibody titers below the seroconversion threshold (defined as a signal threshold >1 at the 1:100 dilution for any isotype) were assigned a value of 25 for graphical representation only. The y-axis is plotted in logarithmic scale.

**Appendix Figure 3. Anti–SARS-CoV-2 serum antibody titers by isotype and detection of SARS-CoV-2 by BinaxNOW™ rapid antigen test, real-time reverse transcription polymerase chain reaction, and viral culture post-diagnosis, by participant (N=11) in a nursing home cohort—Georgia, October 2020–April 2021**

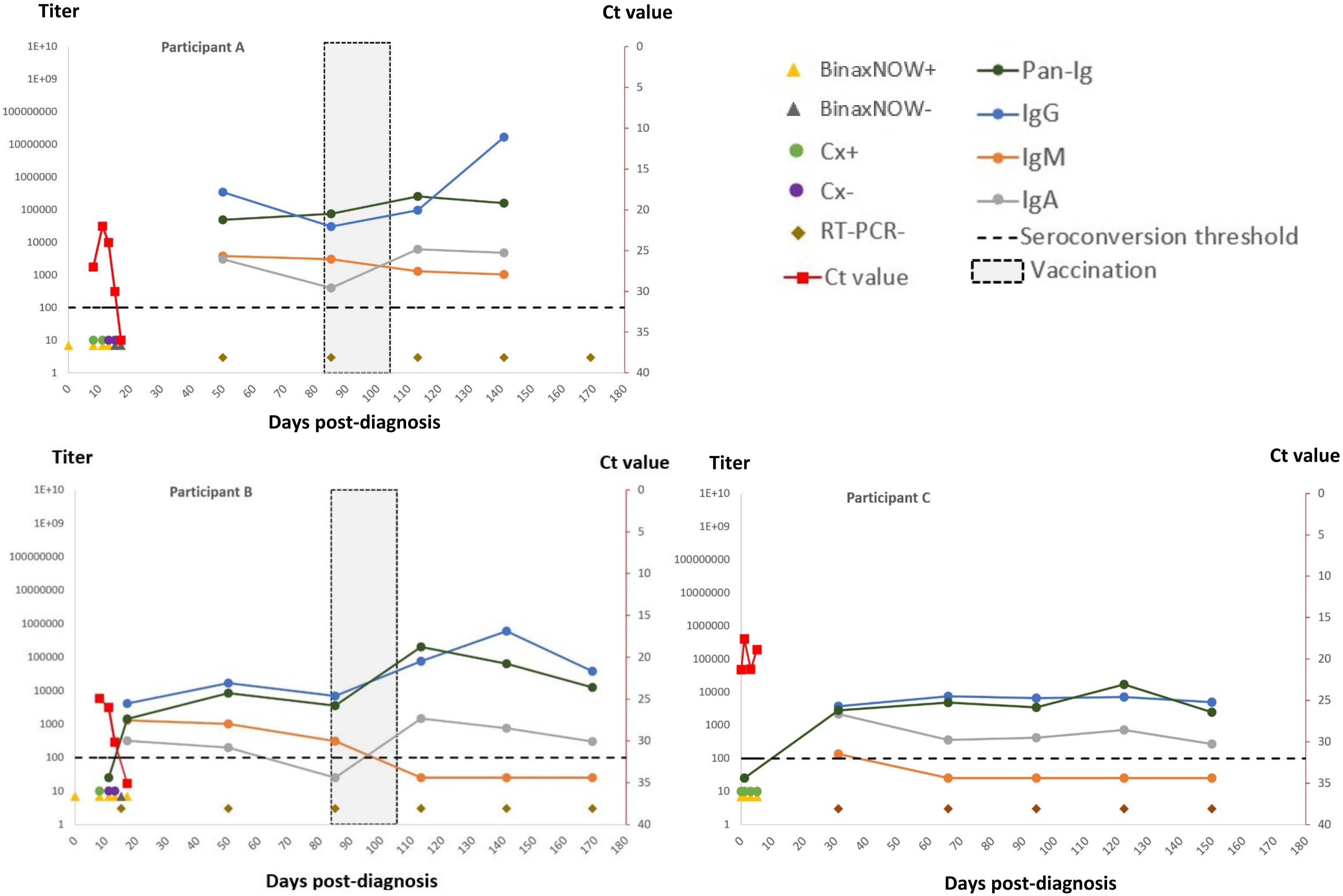

Titer
Ct value
Participant A
Participant B
Participant C
Days post-diagnosis
1E+10
1E+09
100000000
10000000
1000000
100000
10000
1000
100
10
1
0
5
10
15
20
25
30
35
40
0 10 20 30 40 50 60 70 80 90 100 110 120 130 140 150 160 170 180
BinaxNOW+
BinaxNOW-
Cx+
Cx-
RT-PCR-
Ct value
Pan-Ig
IgG
IgM
IgA
Seroconversion threshold
Vaccination

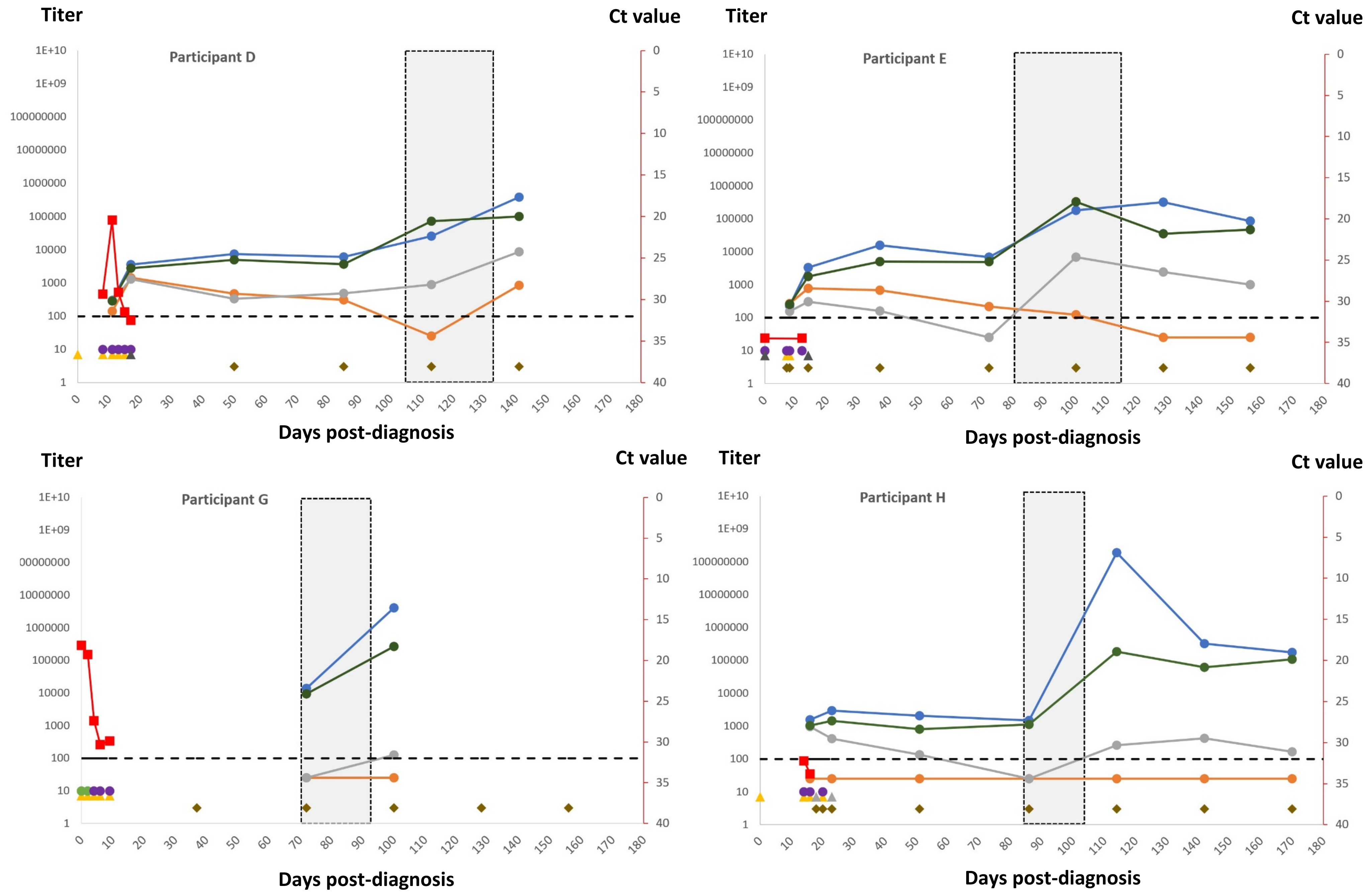

Titer
Ct value
Participant D
Participant E
Participant G
Participant H
Days post-diagnosis

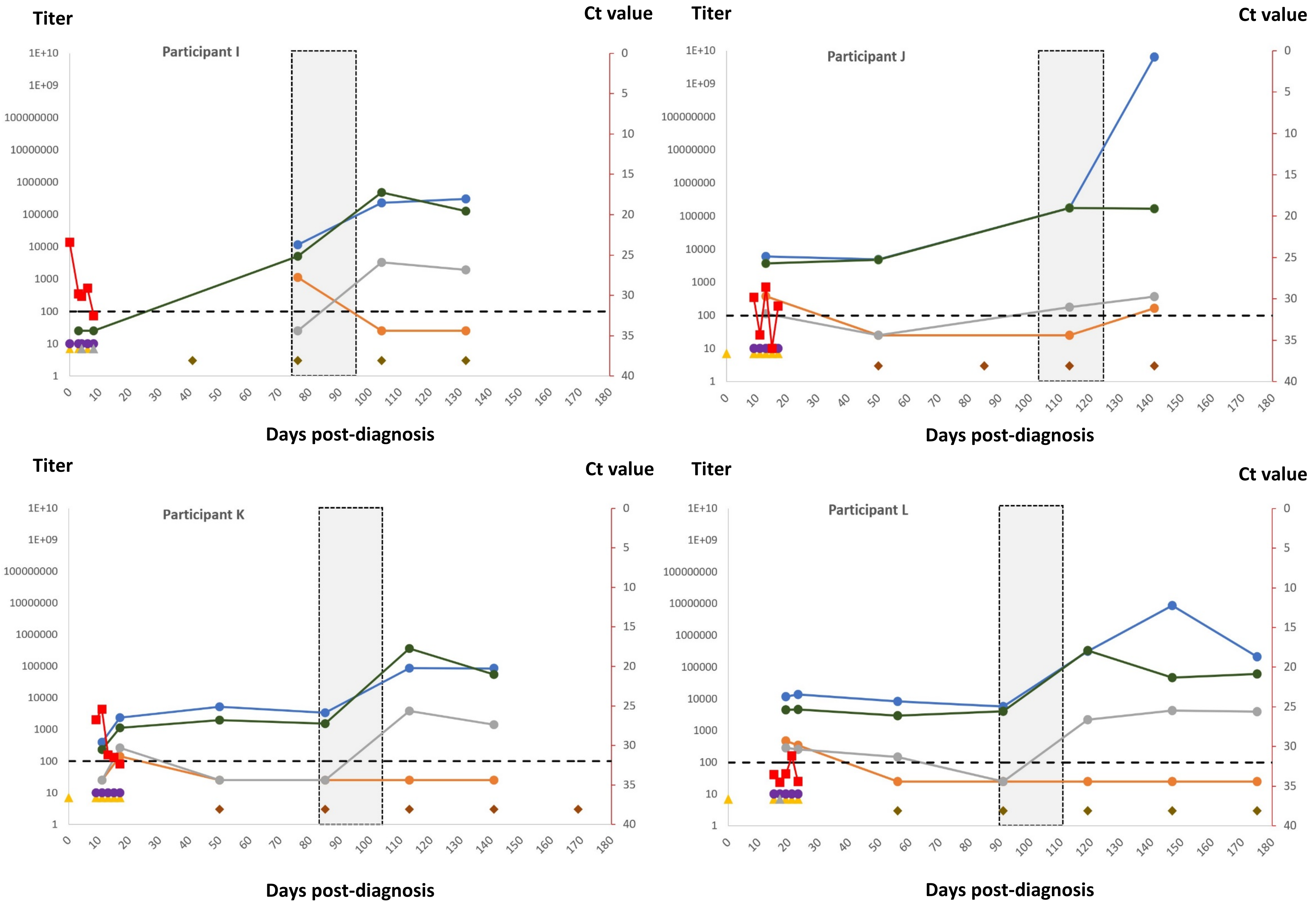
Titer
Ct value
Participant I
Days post-diagnosis
Titer
Ct value
Participant J
Days post-diagnosis
Titer
Ct value
Participant K
Days post-diagnosis
Titer
Ct value
Participant L
Days post-diagnosis
1E+10
1E+09
100000000
10000000
1000000
100000
10000
1000
100
10
1
0
5
10
15
20
25
30
35
40
0
10
20
30
40
50
60
70
80
90
100
110
120
130
140
150
160
170
180

Abbreviations: BinaxNOW = BinaxNOW™ COVID-19 Ag Card; RT-PCR = real-time reverse transcription polymerase chain reaction; Cx = viral culture

Blood samples were obtained at several timepoints between 1–150 days post-diagnosis (time period after each participant's first positive SARS-CoV-2 test result). Due to challenges phlebotomizing some patients, serum titers were not generated for each participant for each visit. Pfizer-BioNTech COVID-19 vaccines were administered at the facility between 71–105 days post-diagnosis; participant C declined vaccination. Participants with serum antibody titers below the seroconversion threshold (defined as a signal threshold >1 at the 1:100 dilution for any isotype) were assigned a value of 25 for graphical representation. The y-axis is plotted in logarithmic scale.
